# Evaluation of Large Language Models in the Clinical Management of Patients With Upper Gastrointestinal Bleeding : Insights From Real-World Patient Data

**DOI:** 10.1101/2025.11.10.25339858

**Authors:** Mohsen Rajabnia, Farbod Davoodi, Mahsa Hajisafarali, Shadi Asadinia, Mahsa Shiri, Fatemeh Shirzad, Peyman Saeedi, Mahsa Mohammadi

## Abstract

**Objective:** Upper gastrointestinal bleeding (UGIB) is a life-threatening emergency requiring rapid risk assessment. Current scoring tools have limited accuracy. Large language models (LLMs) may support clinical decision-making, but their role in UGIB management is unclear. This study evaluated LLMs for patient risk classification, prediction of endoscopic findings, and alignment with routine clinical decision-making.

**Methods:** In this retrospective study, we analyzed electronic health records(EHRs) of 384 UGIB patients presented to two referral centers in Karaj, Iran, between March and December 2024. Included cases underwent upper gastrointestinal endoscopy; incomplete records were excluded. Five LLMs including GPT-5, Llama 4, Gemini-2.5-Flash, DeepSeek R1, and Grok were assessed using in-context learning for (i) risk classification, (ii) prediction of probable endoscopic findings, and (iii) clinical justification generation. Performance metrics included accuracy, precision, recall, and F1-score, compared with conventional machine learning models. Two gastroenterologists independently assessed justifications across seven domains: relevance, clarity, originality, completeness, specificity, correctness, and consistency.

**Results:** All LLMs outperformed conventional models (highest baseline accuracy 0.54). GPT-5 achieved the highest risk classification accuracy (0.66), followed by Llama 4 (0.64). Grok performed best in predicting endoscopic findings (0.32). gastroenterologists noted variability in reasoning: GPT-5 and Grok provided the most complete justifications, though GPT-5 occasionally over-classified urgent cases. Llama-4 and Gemini-2.5-Flash were less specific, while DeepSeek R1 offered detailed patient summaries but lacked predictive outputs.

**Conclusions:** LLMs improved UGIB risk prediction and generate interpretive reasoning, but accuracy limitations, inconsistent reasoning, and occasional risk misclassification highlight the need for clinician oversight and prospective validation before clinical use.

**Key Messages:** *What is already known on this topic:* UGIB is a medical emergency requiring rapid risk stratification and timely management. LLMs are promising tools for clinical decision support, but their role in UGIB management remains unclear.

*What this study adds:* LLMs can improve risk prediction and interpretive reasoning in UGIB, but limitations in accuracy, inconsistent reasoning, and occasional misclassification highlight the need for clinician oversight and prospective validation.

*How this study might affect research, practice, or policy:* LLMs provide structured, human-readable explanations that could support clinical decision-making, potentially reduce unnecessary emergency endoscopies, improve care efficiency, and alleviate physician workload.

## INTRODUCTION

Gastrointestinal bleeding(GIB) is a common medical emergency that requires rapid risk stratification and timely management. UGIB accounts for over half of all GIB cases and is associated with mortality rates ranging from 2% to 10.(1). Urgent endoscopy is often performed for suspected GIB during nights or holidays; however, in many cases, bleeding has already stopped spontaneously by the time of the procedure. Current guidelines emphasize the importance of classifying patients into high-risk groups requiring prompt intervention such as transfusions or hemostatic procedures (endoscopic, surgical, or interventional radiology) and low-risk groups who do not need emergent intervention and can be safely discharged from the emergency department for outpatient management (1, 2). Several scoring systems have been developed and validated to predict both risk and mortality. These include the Rockall score, Glasgow-Blatchford Score (GBS), modified GBS, and AIMS65 score(3). Effective risk stratification can reduce unnecessary emergency endoscopies, thereby improving the efficiency of medical care and alleviating the workload on physicians(1). but the predictive performance of existing tools remains suboptimal and their application in routine clinical practice remains limited(2).

The rapid advancement of LLMs has driven their widespread use and demonstrated their ability to comprehend and generate complex medical information (4-6). Physicians are increasingly integrating these tools into clinical practice. Evidence suggests that, relative to human specialists, LLMs possess a broad understanding across multiple medical domains, underscoring their potential as versatile diagnostic aids (4). Conventional risk scores often fail to capture the complex interplay of clinical, laboratory, and endoscopic factors influencing outcomes. LLMs are simple to implement, integrate diverse data, and provide context-specific predictions with accuracy comparable to complex rule-based approaches. These features make them promising tools for free-text classification in clinical decision support(7, 8).

Recent studies highlight that LLMs can play a valuable role in gastroenterology, particularly in detecting and characterizing GI bleeding. An LLM-based pipeline trained on over 17,000 nursing notes accurately identified overt and recurrent bleeding, improving coding accuracy and achieving an AUC of 0.986 for recurrence(7). Another study compared LLMs with physicians in 67 complex GI cases found that Claude 3.5 Sonnet achieved the highest diagnostic coverage (76.1%), significantly outperforming gastroenterologists and those using traditional resources (4). ChatGPT was also assessed for accuracy in gastroenterology using 16 dialogue-based and 13 definition-based queries reviewed by three gastroenterologists in six quality domains. 44% and 69% of responses, respectively, were appropriate. All outputs were written at a college level but limited readability.(5). Another study assessed the reliability of ChatGPT in answering patient questions about common endoscopic procedures. It gave mostly accurate guidance for routine procedures (like esophagogastroduodenoscopy and colonoscopy) but produced multiple errors for complex interventions(like endoscopic retrograde cholangiopancreatography (ERCP), and endoscopic ultrasound (EUS)) procedures (9). In an effort a domain-specific LLM, GutGPT, was trained on over 190,000 GI disease question–answer datasets derived from real-world physician–patient conversations, medical guidelines, knowledge graphs, and Chinese medical licensing examinations. Compared with 16 existing models, GutGPT demonstrated superior performance, achieving a 9.6% improvement in diagnostic accuracy in expert assessments and a 22.5% on public datasets, while also providing empathetic responses, highlighting its potential as a decision-support and patient engagement tool(10).

Together, these findings underscore the potential of LLMs as effective tools for both clinical decision support and the enhancement of administrative processes in gastroenterology. However, their role in the management of UGIB has not yet been systematically evaluated. Considering important limitations in accuracy, reliability, and accessibility that must be addressed before widespread clinical integration. The aim of this study was to evaluate the potential of LLMs to classify patients presenting with UGIB as high-risk requiring urgent intervention or low-risk suitable for outpatient management, and to predict probable endoscopic findings. Additionally, the study assessed the concordance of LLM-based predictions with routine clinical decision-making, as evaluated by two gastroenterologists, to determine their potential as decision-support tools in the acute care setting.

## METHODS

### Study Design

This retrospective observational study was conducted at two referral centers affiliated with Alborz University of Medical Sciences in Karaj, Iran, between March 2024 and December 2024. Both centers provide 24-hour endoscopic and critical care services. The study was approved by the Institutional Ethics Committee (ethical code: IR.ABZUMS.REC.1403.202), and written informed consent was obtained from all participants. The study population included patients presenting to the emergency department with suspected UGIB defined as hematemesis, coffee-ground emesis, melena, or hematochezia of presumed upper gastrointestinal origin. Patients were eligible if they underwent upper gastrointestinal endoscopy during hospitalization. Those without endoscopy or with incomplete clinical records were excluded.

From each EHR, a standardized dataset was collected, including demographics, presenting symptoms, comorbidities, lifestyle and medication history, vital signs and physical examination at admission, relevant laboratory investigations, and endoscopic findings. Data were collected independently by four researchers, and discrepancies were resolved by consensus. A total of 384 cases met the eligibility criteria and were included in the final dataset. All data were entered into a structured Microsoft Excel spreadsheet and checked for accuracy, completeness, and consistency prior to analysis. The collected variables for each patient are summarized in Table 1.

**Table 1.** Variables collected from EHRs of patients with UGIB.

| Domain | Variable | Description |
| --- | --- | --- |
| Demographics | Age | Recorded in years |
|  | Sex | Male / Female |
| Presenting features | Melena | Presence of black, tarry stools (Yes/No) |
|  | Hematemesis | Vomiting of blood (Yes/No) |
|  | Syncope | Transient loss of consciousness (Yes/No) |
|  | Shock | Hemodynamic instability at presentation (Yes/No) |
| **Comorbidities** | Diabetes mellitus | Documented diagnosis (Yes/No) |
|  | Hypertension | Documented diagnosis (Yes/No) |
|  | Cardiovascular disease | Ischemic heart disease, heart failure, or arrhythmia (Yes/No) |
|  | Deep vein thrombosis | History of DVT (Yes/No) |
|  | Chronic obstructive pulmonary disease | Documented COPD (Yes/No) |
|  | Pulmonary embolism | History of PE (Yes/No) |
|  | Chronic liver disease | Non-cirrhotic liver disease (Yes/No) |
|  | Cirrhosis | Confirmed diagnosis of cirrhosis (Yes/No) |
|  | Chronic renal disease | Chronic kidney disease, stages 3–5 (Yes/No) |
|  | Anemia | Pre-existing anemia (Yes/No) |
| **Lifestyle & medications** | Smoking | Active tobacco use (Yes/No) |
|  | Alcohol use | Documented alcohol consumption (Yes/No) |
|  | Peptic ulcer disease | Past history of PUD (Yes/No) |
|  | Previous GI bleeding | History of UGIB or LGIB (Yes/No) |
|  | Anti-acid therapy | Proton pump inhibitors or H2-blockers (Yes/No) |
|  | NSAID use | Non-steroidal anti-inflammatory drugs (Yes/No) |
|  | Anticoagulant use | Warfarin, DOACs, or heparin (Yes/No) |
| **Clinical parameters** | Mental status | Normal / Altered |
|  | Blood pressure | Measured in mmHg |
|  | Heart rate | Beats per minute |
| **Laboratory values** | Hemoglobin (Hb) | g/dL (at admission, 6h, 12h) |
|  | Albumin | g/dL |
|  | Blood urea nitrogen (BUN) | mg/dL |
|  | Creatinine | mg/dL |
|  | C-reactive protein (CRP) | mg/L |
| | Platelet count | $\times 1000 /\mu\text{L}$ |
|  | Prothrombin time (PT) | Seconds |
|  | Partial thromboplastin time (PTT) | Seconds |
|  | International normalized ratio (INR) | Ratio |
| **Endoscopy** | Endoscopic findings | Endoscopic findings, including lesion type and, when a peptic ulcer was present, the corresponding Forrest classification |

### Model Evaluation

We first evaluated conventional ML models to establish baseline performance for prediction tasks.

For evaluation of LLMs, a structured prompt incorporating relevant clinical and laboratory data for each case was generated. We applied in-context-learning (11) techniques, including chain-of-thought (12) and tree of thought(13) to guide the models’ stepwise evaluation of clinical parameters and decision pathways. Prompts first presented the patient’s symptoms and then requested an assessment of the risk of requiring endoscopic intervention. To ensure systematic inference, model outputs were structured according to a Tree-of-Thought framework. Each prompt elicited three prespecified outputs:

1. Endoscopy timing: classification of patients as high-risk requiring urgent intervention or low-risk eligible for outpatient management.
2. Predicted endoscopic findings: including peptic ulcers (Forrest I–III), erosive gastropathy, esophageal varices, Mallory–Weiss tears, tumors, and other lesions.
3. Rationale: explanation of the reasoning behind the model’s predictions.

We initially evaluated several LLMs including MedGemma(14), MedAlpaca(15), Meditron(16), MedicalBERT(17), ClinicalBERT(18), and BioBERT(18) before selecting GPT-5(19), Gemini-2.5-Flash(20), Llama 4(21), DeepSeek R1(22, 23), and Grok(24). We excluded the former set for several reasons. First, models such as MedAlpaca, Meditron, and BERT-based variants have limited context windows, which is insufficient to process our full clinical prompts. Second, because these models generally have fewer parameters and narrower domain fine-tuning, they often fail to achieve adequate performance on metrics such as accuracy, precision, recall, and F1-score when applied to complex medical reasoning tasks. Third, some specialized medical models require computational resources that exceed the practical limits of our infrastructure. The selected models supported larger input contexts, produced higher-quality outputs, and were compatible with our computational resources.

Each prompt was submitted independently to the five selected LLMs and the generated responses were recorded, annotated, and evaluated using predefined criteria for accuracy, precision, recall, and F1-score. This approach ensured reproducibility and standardized assessment of model performance across varied clinical scenarios.

## RESULTS

### Application of Conventional Machine Learning Models

We initially assessed conventional ML models to establish baseline performance for the prediction tasks. Table 2 presents accuracy, F1-score, and computation time for each model. Overall, conventional models showed moderate to low predictive performance. LabelSpreading achieved the highest F1-score (0.52), while BaggingClassifier demonstrated balanced accuracy with minimal computation time.

**Table 2.** Comparative Performance Metrics of Conventional ML Models.

| Model | Accuracy | F1-Score |  |
| --- | --- | --- | --- |
| BaggingClassifier | 0.49 | 0.48 | 0.07 |
| LabelSpreading | 0.54 | 0.52 | 0.11 |
| KNeighborsClassifier | 0.49 | 0.46 | 0.05 |
| ExtraTreeClassifier | 0.44 | 0.44 | 0.02 |
| AdaBoostClassifier | 0.32 | 0.21 | 0.18 |
| DecisionTreeClassifier | 0.39 | 0.38 | 0.04 |
| LGBMClassifier | 0.37 | 0.36 | 0.32 |
| BernoulliNB | 0.37 | 0.33 | 0.03 |
| RandomForestClassifier | 0.37 | 0.34 | 0.41 |
| SVC | 0.29 | 0.25 | 0.09 |
| RidgeClassifierCV | 0.29 | 0.26 | 0.06 |
| RidgeClassifier | 0.29 | 0.26 | 0.02 |
| LogisticRegression | 0.29 | 0.26 | 0.04 |
| LinearDiscriminantAnalysis | 0.29 | 0.26 | 0.05 |
| LinearSVC | 0.29 | 0.26 | 0.07 |
| DummyClassifier | 0.24 | 0.10 | 0.04 |
| CalibratedClassifierCV | 0.24 | 0.13 | 0.18 |
| PassiveAggressiveClassifier | 0.24 | 0.23 | 0.04 |
| NearestCentroid | 0.12 | 0.14 | 0.05 |
| SGDClassifier | 0.20 | 0.18 | 0.13 |
| Perceptron | 0.20 | 0.17 | 0.02 |
| GaussianNB | 0.12 | 0.17 | 0.02 |

### LLM Performance in Risk Stratification

Table 3 summarizes the performance of evaluated LLMs in binary classifying patients into high-risk and low-risk groups. GPT-5 and Llama 4 achieved the highest overall accuracy (0.66 and 0.64, respectively), indicating robust and balanced performance. Grok and DeepSeek R1 demonstrated moderate performance, while Gemini-2.5-Flash showed the lowest overall accuracy (0.40) but high recall for high-risk cases (0.89), indicating sensitivity at the expense of specificity.

**Table 3.**
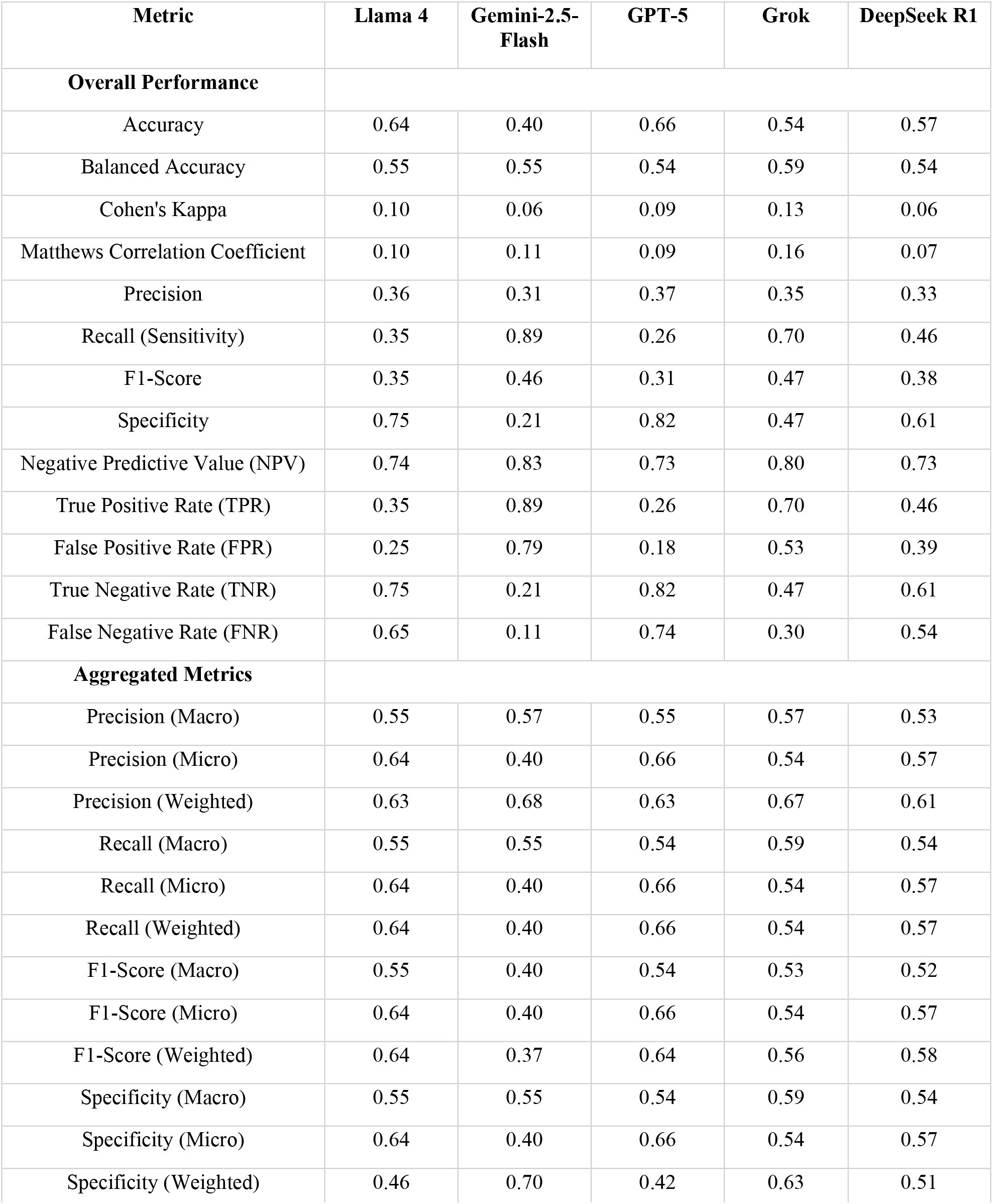
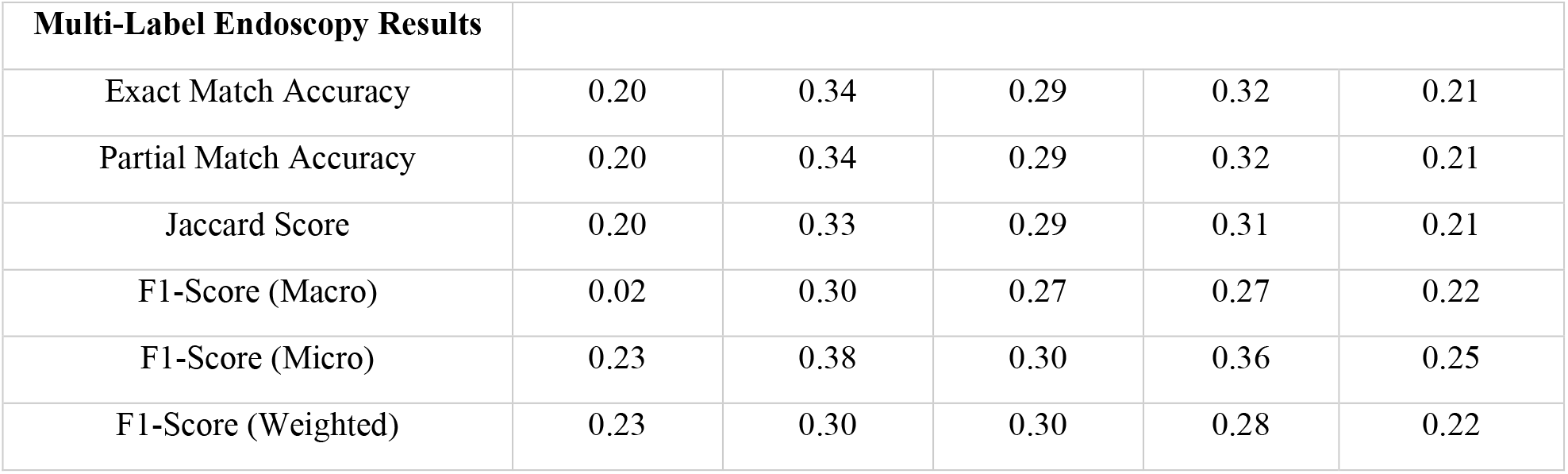
Performance of LLMs in classifying Patients into High- and Low-Risk Groups and Multi-label prediction of endoscopic findings.

| Metric | Llama 4 | Gemini-2.5-Flash | GPT-5 | Grok | DeepSeek R1 |
| --- | --- | --- | --- | --- | --- |
| Overall Performance |  |  |  |  |  |
| Accuracy | 0.64 | 0.40 | 0.66 | 0.54 | 0.57 |
| Balanced Accuracy | 0.55 | 0.55 | 0.54 | 0.59 | 0.54 |
| Cohen's Kappa | 0.10 | 0.06 | 0.09 | 0.13 | 0.06 |
| Matthews Correlation Coefficient | 0.10 | 0.11 | 0.09 | 0.16 | 0.07 |
| Precision | 0.36 | 0.31 | 0.37 | 0.35 | 0.33 |
| Recall (Sensitivity) | 0.35 | 0.89 | 0.26 | 0.70 | 0.46 |
| F1-Score | 0.35 | 0.46 | 0.31 | 0.47 | 0.38 |
| Specificity | 0.75 | 0.21 | 0.82 | 0.47 | 0.61 |
| Negative Predictive Value (NPV) | 0.74 | 0.83 | 0.73 | 0.80 | 0.73 |
| True Positive Rate (TPR) | 0.35 | 0.89 | 0.26 | 0.70 | 0.46 |
| False Positive Rate (FPR) | 0.25 | 0.79 | 0.18 | 0.53 | 0.39 |
| True Negative Rate (TNR) | 0.75 | 0.21 | 0.82 | 0.47 | 0.61 |
| False Negative Rate (FNR) | 0.65 | 0.11 | 0.74 | 0.30 | 0.54 |
| Aggregated Metrics |  |  |  |  |  |
| Precision (Macro) | 0.55 | 0.57 | 0.55 | 0.57 | 0.53 |
| Precision (Micro) | 0.64 | 0.40 | 0.66 | 0.54 | 0.57 |
| Precision (Weighted) | 0.63 | 0.68 | 0.63 | 0.67 | 0.61 |
| Recall (Macro) | 0.55 | 0.55 | 0.54 | 0.59 | 0.54 |
| Recall (Micro) | 0.64 | 0.40 | 0.66 | 0.54 | 0.57 |
| Recall (Weighted) | 0.64 | 0.40 | 0.66 | 0.54 | 0.57 |
| F1-Score (Macro) | 0.55 | 0.40 | 0.54 | 0.53 | 0.52 |
| F1-Score (Micro) | 0.64 | 0.40 | 0.66 | 0.54 | 0.57 |
| F1-Score (Weighted) | 0.64 | 0.37 | 0.64 | 0.56 | 0.58 |
| Specificity (Macro) | 0.55 | 0.55 | 0.54 | 0.59 | 0.54 |
| Specificity (Micro) | 0.64 | 0.40 | 0.66 | 0.54 | 0.57 |
| Specificity (Weighted) | 0.46 | 0.70 | 0.42 | 0.63 | 0.51 |

| Multi-Label Endoscopy Results |  |  |  |  |  |
| --- | --- | --- | --- | --- | --- |
| Exact Match Accuracy | 0.20 | 0.34 | 0.29 | 0.32 | 0.21 |
| Partial Match Accuracy | 0.20 | 0.34 | 0.29 | 0.32 | 0.21 |
| Jaccard Score | 0.20 | 0.33 | 0.29 | 0.31 | 0.21 |
| F1-Score (Macro) | 0.02 | 0.30 | 0.27 | 0.27 | 0.22 |
| F1-Score (Micro) | 0.23 | 0.38 | 0.30 | 0.36 | 0.25 |
| F1-Score (Weighted) | 0.23 | 0.30 | 0.30 | 0.28 | 0.22 |

Multi-label prediction of endoscopic findings revealed that Grok achieved the highest exact and partial match accuracy (0.32), although F1-scores remained low across all models, reflecting the complexity of multi-label classification. These findings suggest that GPT-5 and Llama 4 are generally well-balanced, while Grok may be preferable for multi-label prediction tasks. Overall performance across accuracy, precision, recall, F1-score, exact match accuracy, and area under the curve (AUC) is presented in Figure 1, highlighting the relative strengths and limitations of each model.

**Figure 1.**
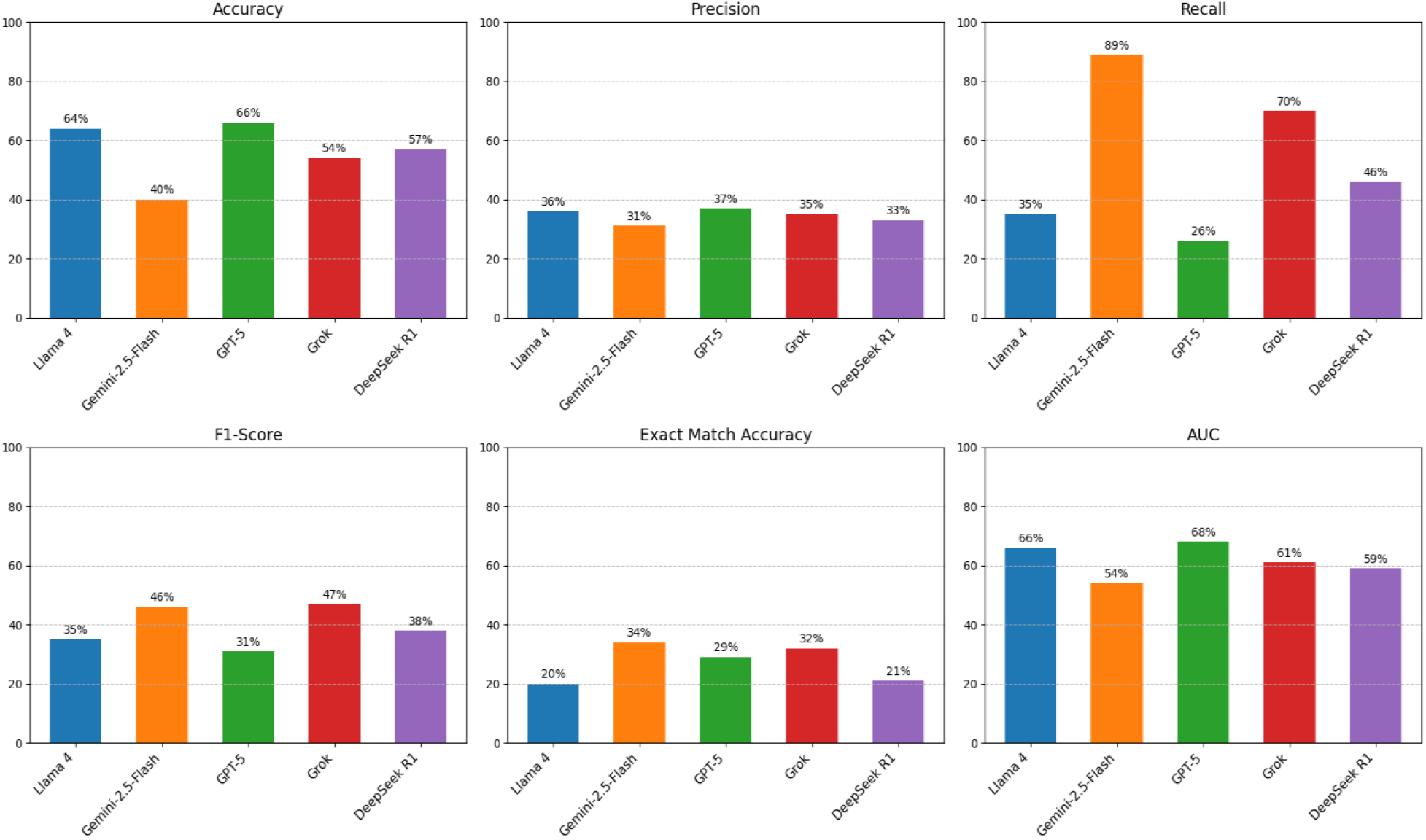
Performance of evaluated large language models across key metrics, including accuracy, precision, recall, F1-score, exact match accuracy, and AUC.

Model performance in predicting high- and low-risk patients is shown in Table 4. GPT-5 and Llama 4 demonstrated the most balanced performance, with GPT-5 achieving the highest precision for high-risk patients (0.37) and Gemini-2.5-Flash the highest for low-risk patients (0.83). Gemini-2.5-Flash had high recall for high-risk patients (0.89) but low recall for low-risk patients (0.21), indicating a tendency to over-predict high-risk cases. Grok and DeepSeek R1 showed moderate, balanced performance. Specificity was generally higher for low-risk predictions, while F1-scores highlight the challenge of achieving strong performance across both classes. Overall, low-risk patients were consistently identified with higher precision, recall, and F1-scores than high-risk patients, indicating that the models were generally more effective at detecting low-risk cases.

**Table 4.**
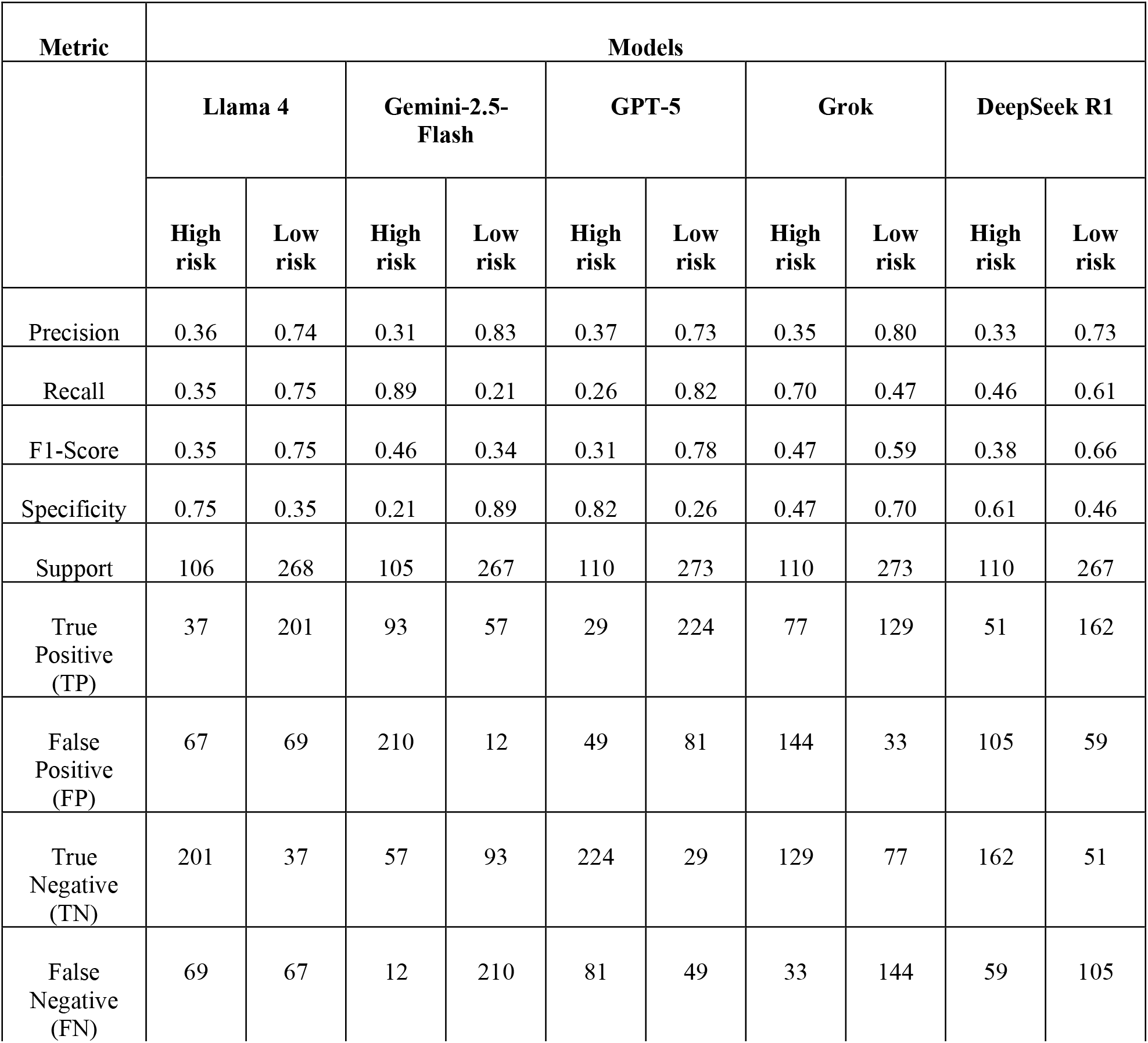
Binary Classification Performance of ML Models in Predicting High-Risk and Low-Risk Patients.

| Metric | Models |  |  |  |  |  |  |  |  |  |
| --- | --- | --- | --- | --- | --- | --- | --- | --- | --- | --- |
|  | Llama 4 |  | Gemini-2.5-Flash |  | GPT-5 |  | Grok |  | DeepSeek R1 |  |
|  | High risk | Low risk | High risk | Low risk | High risk | Low risk | High risk | Low risk | High risk | Low risk |
| Precision | 0.36 | 0.74 | 0.31 | 0.83 | 0.37 | 0.73 | 0.35 | 0.80 | 0.33 | 0.73 |
| Recall | 0.35 | 0.75 | 0.89 | 0.21 | 0.26 | 0.82 | 0.70 | 0.47 | 0.46 | 0.61 |
| F1-Score | 0.35 | 0.75 | 0.46 | 0.34 | 0.31 | 0.78 | 0.47 | 0.59 | 0.38 | 0.66 |
| Specificity | 0.75 | 0.35 | 0.21 | 0.89 | 0.82 | 0.26 | 0.47 | 0.70 | 0.61 | 0.46 |
| Support | 106 | 268 | 105 | 267 | 110 | 273 | 110 | 273 | 110 | 267 |
| True Positive (TP) | 37 | 201 | 93 | 57 | 29 | 224 | 77 | 129 | 51 | 162 |
| False Positive (FP) | 67 | 69 | 210 | 12 | 49 | 81 | 144 | 33 | 105 | 59 |
| True Negative (TN) | 201 | 37 | 57 | 93 | 224 | 29 | 129 | 77 | 162 | 51 |
| False Negative (FN) | 69 | 67 | 12 | 210 | 81 | 49 | 33 | 144 | 59 | 105 |

### Human Justification Evaluation Results

by two expert gastroenterologists assessed a randomly selected sample of 100 datas to evaluate The quality and Clinical reliability of justifications produced by Llama-4, GPT-5, Gemini-2.5-Flash, Grok, and DeepSeek R1. Justifications were evaluated across seven metrics:

1. Relevance – alignment of the justification with the patient’s health record.
2. Clarity – readability and comprehensibility of the text.
3. Originality – evidence of synthesis rather than verbatim reproduction.
4. Completeness – coverage of all relevant aspects of the patient’s record.
5. Specificity – precision in addressing the prediction.
6. Correctness – factual accuracy and consistency with clinical information.
7. Consistency – logical coherence and alignment with documented reasoning.

Strong concordance between human and automated assessments supported the reliability of the evaluation framework.

Model-specific observations included:

- GPT-5: produced complete but occasionally repetitive justifications, relying on key criteria and over-classifying urgent cases when GIB co-occurred with tachycardia.
- Llama-4: generated short, vague, and low-specificity justifications; occasionally correct but generally weak in reasoning.
- Grok: provided more complete explanations with sometimes appropriate decisions, though reasoning was occasionally inconsistent or incoherent.
- Gemini-2.5-Flash: integrated multiple clinical features in structured justifications but frequently misclassified cases due to incorrect prioritization or overly general reasoning.
- DeepSeek R1: provided detailed, patient-specific summaries describing clinical features, but did not generate predictive results, and showed occasional tendencies toward overtreatment.

Overall, the Human Justification Evaluation of models demonstrated variation between justification depth, reasoning consistency, and predictive accuracy.

## DISCUSSION

In this study, we evaluated different LLMs for predicting endoscopic findings and classifying patients with UGIB into high-risk requiring urgent intervention or low-risk eligible for outpatient management using in-context-learning, chain-of-thought, and tree-of-thought techniques for prompting. We selected five LLMs—GPT-5, Llama 4, Grok, DeepSeek R1, and Gemini-2.5-Flash—based on performance and feasibility. Among the selected models, GPT-5 and Llama 4 demonstrated moderate accuracy with balanced precision, recall, and F1-scores, while Grok performed best for multi-label prediction of endoscopic findings. Conventional ML models showed limited predictive ability, highlighting the advantage of LLMs in integrating complex clinical, laboratory, and procedural data.

These selected LLMs generated structured, human-readable explanations, advancing explainable AI in gastroenterology but Models outputs varied notably. GPT-5 produced the most complete justifications but was sometimes repetitive and over-classified urgent cases, while Grok offered similarly detailed explanations with less consistent reasoning. Llama-4 generated brief, vague outputs, and Gemini-2.5-Flash integrated multiple clinical features but often misclassified cases. DeepSeek R1 excelled in patient-specific detail but did not provide predictive outputs and occasionally suggested overtreatment. Overall, GPT-5 and Grok showed stronger reasoning, Llama-4 and Gemini-2.5-Flash were limited in specificity and accuracy, and DeepSeek R1 was descriptive but non-predictive.

Our findings align with previous studies demonstrating the potential of LLMs to support complex medical reasoning in gastroenterology(4, 5, 7, 9, 10). By synthesizing diverse clinical features, LLMs may overcome limitations of traditional scoring systems, which are often rigid and underperform in heterogeneous populations. Nonetheless, modest overall predictive accuracy and multi-label performance reveal ongoing challenges, including handling rare presentations, heterogeneous datasets, and avoiding mis-triage of high-risk patients.

Limitations of this study include its retrospective design and reliance on medical records, which may be incomplete or inconsistently documented, limiting causal inference. Only patients undergoing endoscopy were included, potentially excluding milder cases and introducing selection bias. The study was conducted at two referral centers in a single region over a limited period, restricting generalizability. Automated LLM-based evaluation processes may inflate performance metrics or reveal inconsistencies between models, reflecting the trade-off between scalability and nuanced clinical assessment. Models like MedAlpaca, Meditron, and BERT variants were excluded due to limited context, narrower domain tuning, and high computational demands.

Ethical considerations are essential when applying LLMs in clinical practice. Models trained on large datasets may contain inherent biases—legal, technical, or linguistic—which can influence generated explanations and decision-making, potentially compromising fairness and accuracy. LLMs may also produce unreal or factually incorrect outputs, potentially leading to misinterpretation of patient records. Automated predictions should therefore complement, not replace, human expertise, with clinicians remaining central to verifying and interpreting AI-generated insights.

Future research should focus on prospective validation, integration with electronic health records, domain-specific fine-tuning, and evaluation of effects on patient outcomes, workflow efficiency, and healthcare resource utilization.

In conclusion, LLMs demonstrate promising potential for risk stratification and endoscopic prediction in patients with UGIB, offering structured, interpretable explanations that surpass traditional scoring systems in handling complex clinical data. All models, however, exhibited limitations in accuracy, reasoning consistency, and risk misclassification. While these findings support the complementary use of LLMs in gastroenterology, human expertise remains essential, and further prospective validation and domain-specific optimization are needed to ensure safe and effective clinical implementation.

## Data Availability

All data produced in the present study are available upon reasonable request to the authors

## ACKNOWLEDGEMENTS

The authors gratefully acknowledge the support of the Cardiovascular Research Center (CRC) at Alborz University of Medical Sciences and Imam Ali Hospital, Karaj, Iran. We also thank the study participants for providing their data. The authors thank Dr Bahar Seifi for her valuable assistance in the preparation of this manuscript.

## AI use declaration

ChatGPT (OpenAI, GPT-5) was used solely for language editing, including improving grammar, clarity, and style. The authors reviewed and approved all AI-generated suggestions, and the final content reflects their own interpretation and conclusions.

